# Provider-Level and Specialty Variation in ARNI and SGLT2i Use After Heart Failure Hospitalization

**DOI:** 10.64898/2026.11.06.26367478

**Authors:** Taufiq Salahuddin, Annika Patten, Christine Chow, Paul L. Hess, Larry A. Allen, Carol Simons, Utibe R. Essien, Fatima Rodriguez, Laura Scherer, Chelsea Leonard, Daniel Matlock, P. Michael Ho, Christian Helfrich, Jacob A. Doll

## Abstract

**Background:** Angiotensin receptor-neprilysin inhibitors (ARNI) and sodium-glucose cotransporter-2 inhibitors (SGLT2i) are incompletely utilized in heart failure with reduced ejection fraction (HFrEF). The degree to which use varies at the individual level and within specialties is unknown. We aimed to characterize provider-level and specialty-level variation in ARNI and SGLT2i prescribing after HF hospitalization.

**Methods:** This retrospective cohort study included Veterans Affairs (VA) providers seeing patients within 6 months of HFrEF hospitalization 2017-2023. We used descriptive statistics and site-adjusted multilevel logistic regression to examine prescribing by medical specialty, provider type, and provider sex. Adjusted median odds ratios (mOR) and empirical (unadjusted) provider-level rate ratios (RRs) by percentile were used to describe provider-level variation. Clinical pharmacy visits were assessed as an independent factor and as an interaction with specialty.

**Results:** Among 7,946 providers caring for 76,795 patients across 211,267 outpatient encounters, provider-level variation remained substantial after adjustment for patient characteristics and site-level clustering (mOR 1.45 for ARNI; 1.46 for SGLT2i). Within every specialty, providers ranged from near- zero to near-100% use. Relative to the median provider in the same specialty, RRs ranged from 0-0.31 (5th percentile), 0.37-0.71 (25th), 1.38-1.66 (75th), to 1.8-3.2 (95th). Cardiology visits carried the highest adjusted odds of ARNI (OR 2.30, 95% CI 2.18-2.44) and SGLT2i use (OR 1.99, 95% CI 1.88-2.11) and most new starts. Pharmacist visits were independently associated with greater use of ARNI (OR 1.14, 1.06-1.23) and SGLT2i (OR 1.15, 1.06-1.24), with no effect modification by specialty. In early 2023, 58.4% (ARNI) and 40.2% (SGLT2i) of post-hospitalization HF visits had untreated patients.

**Conclusions:** Provider-level variation in ARNI and SGLT2i use after guideline incorporation was large within every specialty and independent of site effects. Though specific specialties have higher use, individual providers, not specialty alone, may be an important target for efforts to close gaps in therapy.

**CLINICAL PERSPECTIVE:** *What Is New?:*

- In the years following guideline incorporation, provider-level use of ARNI and SGLT2i after heart failure hospitalization ranged from near 0% to near 100% use within every specialty. Providers at the 95th percentile prescribed at 1.8 to 3.2 times the rate the median provider of the same specialty.
- Provider-level variation persisted after accounting for patient characteristics and site-level clustering (median odds ratios 1.45 for ARNI and 1.46 for SGLT2i) in a health system with a national formulary and low copayments.

*What Are the Clinical Implications?:*

- Individual practice patterns within every specialty may be an important target for efforts to close gaps in guideline-directed therapy, and the drivers of this variation warrant study.
- Visits with cardiology and endocrinology providers were associated with greatest use of both medications, supporting specialty care for HF. Clinical pharmacists could play an important adjunctive role.

## INTRODUCTION

Over the last decade, a new generation of medical therapies for heart failure with reduced ejection fraction (HFrEF) has emerged with demonstrated reductions in mortality rates and HF admissions.^1–6^ Despite evidence to support “quadruple therapy” including renin-angiotensin-aldosterone inhibitors (with emphasis on angiotensin receptor-neprilysin inhibitors [ARNI]), sodium glucose co-transporter 2 inhibitors (SGLT2i), beta-blockers, and mineralocorticoid receptor antagonists (MRA),^7^ achievement of this medical therapy goal for HFrEF in routine clinical practice remains low.^8–13^ One recent analysis estimated that 82% of patients were eligible for quadruple therapy, but only 15.3% achieved it.^14^ The greatest opportunities for improvement were noted for ARNI (31.5% use at hospital discharge) and SGLT2i (26.1%), potentially due to the more recent approval, access challenges, and high cost of these medications compared with other indicated classes.^15–18^ Rapid initiation and up-titration of HF GDMT medications during and after HF hospitalization was proven safe and effective in the STRONG HF randomized trial,^19^ with subsequent calls for post-discharge care delivered by multidisciplinary HF teams. However, despite some prior analyses, there is still limited understanding of how individual provider- level characteristics, including provider type (Physician vs Advance Practice Provider training) and specialty are associated with use of ARNI and SGLT2i in the US.^20–23^

The national integrated VA healthcare system may serve as an ideal setting to study provider use of novel medications. VA has a national formulary and low out-of-pocket costs, standardizing access to branded medications for most patients.^24^ Despite this, prior reports from VA have shown substantial site- level variation in ARNI and SGLT2i use.^25–27^ We aimed to characterize provider-level variation in ARNI and SGLT2i utilization and identify potential opportunities for improvement in achievement of optimal HF care related to provider characteristics. We hypothesized that patients treated by specialists and pharmacists would be more likely to receive ARNIs and SGLT2is. We also measured the variation among individual providers within each specialty, providing a snapshot of provider-level adoption in the years after guideline incorporation of these therapies.

## METHODS

### Study Cohort

This was an exploratory, retrospective cohort study analyzing provider-level medication use in outpatient encounters after HF hospitalization. The cohort was constructed by identifying eligible patient- provider encounters within 6 months of hospitalization for heart failure with reduced ejection fractions (HFrEF) between October 1, 2017 and March 31, 2023 within VA (Fiscal Years 2018-2023). The methodology for identifying Veterans with HFrEF has been previously described.^26^ Patients with an inpatient admission for systolic, combined systolic and diastolic, or biventricular heart failure were identified using ICD-10 codes (listed in the Supplemental Methods).^28^ If a patient had multiple qualifying hospitalizations, one was randomly selected as the index hospitalization since overall levels of medication use have varied over time^26^ and the goal of the analysis was to assess provider-level variation. ARNI use was assessed over the full study period but evaluation of SGLT2i use was limited to the period after October 1^st^, 2019 (VA Fiscal Year 2020) to align with the publication of the DAPA-HF trial establishing the efficacy of SGLT2i for HFrEF. Annual data described in this study corresponds to the VA Fiscal Year (October 1-September 30) unless otherwise stated.

Providers were eligible for inclusion if they had an encounter with an included patient within 6 months following discharge from the index hospitalization. Only providers with at least 5 eligible encounters in the four specialties below were evaluated. For descriptive provider-level analyses, each provider was assigned to the specialty in which they had the most qualifying encounters. Provider characteristics were extracted from VA Informatics and Computing Infrastructure and Corporate Data Warehouse (CDW) and included male or female sex, provider type (physician or Advance Practice Provider [APP: Nurse Practitioner or Physician Assistant/Associate]), and clinical specialty (primary care, cardiology, endocrinology, nephrology). Direct patient care encounters were identified using primary and secondary clinic stop codes; pharmacists and clinical pharmacy visits were identified similarly **(Supplement)**.

### Covariates

To describe patient characteristics that may influence provider use of HF medication, we characterized patient demographics, clinical comorbidities, use of other GDMT classes for heart failure, and laboratory values (e.g. estimated glomerular filtration rate (eGFR) and glycated hemoglobin [HbA1c]). Patient race was self-reported as White, Black or African American, Asian, or Other (American Indian or Alaska native, or Native Hawaiian or Pacific Islander) and ethnicity was characterized as Hispanic/Latino or not Hispanic/Latino based on VA demographic data. Additionally, we obtained VA priority group status as an indicator of out-of-pocket medication costs. Priority groups (1-8) are determined by service history, disability rating, income, and other benefits; Veterans in Priority Group 1 pay no medication copayments, and copayments for all others are capped at $700 per calendar year.^24^

### Outcomes

The primary outcomes were provider-level use rates of 1) ARNI or 2) SGLT2i at an individual patient-provider encounter within 6 months of HFrEF hospitalization. A patient was classified as treated with an ARNI or SGLT2i if the medication was filled within 6 months prior to hospital admission, at hospital discharge, or within 6 months after discharge. This included characterizations of patients already on treatment prior to admission, newly started during or after the hospitalization, or previously trialed and discontinued. If a patient was classified as treated with ARNI or SGLT2i, all providers who had an eligible visit with that patient were attributed as users for that medication class.

### Statistical analysis

Descriptive and summary statistics were used to characterize patients by use or non-use of ARNI and SGLT2i. Medication use was plotted over time by provider characteristics and summarized at the encounter level by specialty for the full study period (2018-2023 for ARNI and 2020-2023 for SGLT2i), with visits in January-March 2023 described separately as the most contemporary data in the cohort. For a more detailed description of medication use patterns by specialty, we further classified medication use as: 1) only before: medication was discontinued during admission or after discharge, 2) continued: patient was already on therapy and it was continued, 3) new start: patient was not on therapy within the 6 months prior to discharge and was started upon discharge or within 6 months of discharge, or 4) untreated: no therapy with the given medication class within 6 months of discharge. To test if providers with high use of ARNI also had high use of SGLT2i, a Pearson Correlation coefficient was calculated.

To further investigate observed differences in medication use by specialty, odds ratios (ORs) were estimated comparing the likelihood of medication use for patients having seen vs not seen each specialty. Odds ratios were adjusted using logistic regression for time of prescription (by fiscal year quarter) as well as patient age, sex, race, hypertension, end-stage renal disease, history of prior myocardial infarction, diabetes, Veteran priority status, and clustering by VA site. Renal disease was represented by end-stage renal disease. This was repeated for 2018 (ARNI), 2021, and 2023 to assess changes over time. Odds ratio 95% confidence intervals are Wald intervals; 95% confidence intervals for the median odds ratios were calculated using non-parametric bootstrapping, resampling providers.

Provider-level variation was summarized with median odds ratios (mOR),^25–27^ the median odds ratio of receiving a prescription when the same patient is seen by two randomly chosen providers, and with each provider’s empirical treatment rate (the proportion of that provider’s qualifying encounters with the medication present). The primary descriptive (unadjusted) measure is the rate ratio: the provider’s medication use rate at a given percentile divided by the median provider rate of the same group (overall or within a specialty); for example, RR of 0.50 describes half the median rate of medication use and RR of 3.0 is three-fold higher rate of medication use. Median odds ratios were adjusted for patient characteristics, including renal function, with random intercepts for VA site and provider.

### Clinical pharmacist analysis

VA clinical pharmacist practitioners practice under facility-level scopes of practice that include prescriptive authority for noncontrolled medications. In VA, pharmacists may prescribe independently or assist other clinical providers with medication initiation and titration,^29^ so we analyzed the association of a pharmacist visit with ARNI and SGLT2i use in two complementary ways: 1) as a distinct provider class assessing independent association with medication use, and 2) as an effect modifier assessing change in association with medication use for a given medical specialty class (i.e. the odds of being prescribed an ARNI when seen by cardiology AND pharmacy vs. cardiology alone), using the logistic regression model described above with inclusion of an interaction term.

All analyses were completed using R version 4.2.1.

### Ethical Approval

This study received ethical oversight and approval from the Colorado Multiple Institutional Review Board (COMIRB), protocol #21-4501. COMIRB granted a waiver of informed consent given the retrospective, observational nature of this research using administrative data with no direct patient contact.

## RESULTS

### Medication use and patient characteristics

Our cohort included 7,946 VA providers with at least 5 qualifying encounters (not including pharmacists) caring for 76,795 unique Veteran patients over 211,267 qualifying clinical encounters from 2018-2023. The SGLT2i analysis was limited to encounters between 2020-2023 and included 5,996 providers caring for 45,298 patients over 121,918 qualifying encounters. In addition, there were 3,583 pharmacists caring for 48,632 patients.

ARNI use increased from 4.9% to 41.6% while SGLT2i use increased from 9.3% to 59.8% over the study period **(Figure S1)**. Patients treated with ARNI and SGLT2i were more likely to be younger, identify as Black, have no medication copayments (Priority group 1), and have lower rates of CKD and dialysis when compared with patients not treated (**Table 1**). Usage rates of other GDMT included 91.6% of patients on beta-blockers and 31.6% on MRAs. Patients treated with ARNI or SGLT2i were also more likely to receive these other GDMT classes. Overall use of angiotensin converting enzyme inhibitors or angiotensin receptor blockers (inclusive of ARNI) was 79.9%.

**Table 1:** Heart Failure (HF) cohort characteristics, stratified by receipt of ARNI and SGLT2i.

|  | ARNI cohort, n = 76,795 |  |  | SGLT2i cohort, n = 45,298 |  |  |
|---|---|---|---|---|---|---|
|  | **No ARNI<br>n=63603** | **ARNI<br>n=13192** | **P-value** | **No SGLT2i<br>n=31630** | **SGLT2i<br>n=13668** | **P-value** |
| Age, mean (SD) | 72.64 (10.26) | 69.61 (10.22) | <0.001 | 73.16 (10.37) | 70.34 (9.68) | <0.001 |
| Male | 62,315 (98.0) | 12,853 (97.4) | <0.001 | 30,929 (97.8) | 13,362 (97.8) | 0.946 |
| Race |  |  | <0.001 |  |  | <0.001 |
| Asian | 286 (0.5) | 77 (0.6) |  | 145 (0.5) | 88 (0.7) |  |
| Black or African American | 14,729 (23.3) | 4,287 (32.8) |  | 7,767 (24.8) | 3,875 (28.7) |  |
| Multiple | 1,340 (2.1) | 268 (2.1) |  | 692 (2.2) | 264 (2.0) |  |
| Other | 833 (1.3) | 173 (1.3) |  | 445 (1.4) | 172 (1.3) |  |
| Unknown | 2,803 (4.4) | 596 (4.6) |  | 1,382 (4.4) | 711 (5.3) |  |
| White | 43,145 (68.3) | 7,657 (58.6) |  | 20,913 (66.7) | 8,407 (62.2) |  |
| Ethnicity |  |  | 0.017 |  |  | <0.001 |
| Hispanic or Latino | 3,571 (5.6) | 823 (6.3) |  | 1,735 (5.5) | 922 (6.8) |  |
| Not Hispanic or Latino | 57,651 (90.8) | 11,863 (90.2) |  | 28,673 (90.9) | 12,176 (89.4) |  |
| Unknown | 2,261 (3.6) | 461 (3.5) |  | 1,132 (3.6) | 522 (3.8) |  |
| Veteran Priority Group 1 | 24,814 (39.0) | 5,346 (40.6) | 0.001 | 12,259 (38.8) | 5,712 (41.8) | <0.001 |
| **Medical Characteristics** |  |  |  |  |  |  |
| BMI, mean (SD) | 29.17 (7.04) | 29.57 (6.89) | <0.001 | 28.76 (6.84) | 29.96 (7.07) | <0.001 |
| Diabetes | 38,397 (60.4) | 7,951 (60.3) | 0.839 | 17,631 (55.7) | 9,978 (73.0) | <0.001 |
| Hypertension | 60,057 (94.4) | 12,496 (94.7) | 0.180 | 29,910 (94.6) | 13,041 (95.4) | <0.001 |
| Chronic Kidney Disease | 31,098 (48.9) | 5,767 (43.7) | <0.001 | 15,740 (49.8) | 6,071 (44.4) | <0.001 |
| Dialysis | 3,584 (5.6) | 276 (2.1) | <0.001 | 2,105 (6.7) | 203 (1.5) | <0.001 |
| COPD | 28,815 (45.3) | 5,251 (39.8) | <0.001 | 14,118 (44.6) | 5,479 (40.1) | <0.001 |
| Peripheral Arterial Disease | 17,568 (27.6) | 3,103 (23.5) | <0.001 | 8,985 (28.4) | 3,454 (25.3) | <0.001 |
| Prior Myocardial Infarction | 21,389 (33.6) | 4,535 (34.4) | 0.101 | 11,079 (35.0) | 4,953 (36.2) | 0.014 |
| Prior PCI | 15,601 (24.5) | 3,132 (23.7) | 0.057 | 7,515 (23.8) | 3,654 (26.7) | <0.001 |
| Prior CABG | 20,329 (32.0) | 3,748 (28.4) | <0.001 | 9,815 (31.0) | 4,362 (31.9) | 0.064 |
| Prior Stroke/TIA | 12,906 (20.3) | 2,414 (18.3) | <0.001 | 6,502 (20.6) | 2,588 (18.9) | <0.001 |
| Tobacco Use | 34,519 (54.3) | 7,392 (56.0) | <0.001 | 17,506 (55.3) | 7,791 (57.0) | 0.001 |
| **Labs, Mean (SD)** |  |  |  |  |  |  |
| eGFR, mL/min/1.73m<sup>2</sup> | 61.42 (31.66) | 64.22 (24.76) | <0.001 | 59.88 (32.14) | 63.99 (23.55) | <0.001 |
| Hemoglobin A1c, % | 7.06 (1.77) | 7.01 (1.76) | 0.053 | 6.80 (1.67) | 7.37 (1.86) | <0.001 |
| **Medical therapy** |  |  |  |  |  |  |
| Beta Blockers | 57,489 (90.4) | 12,841 (97.3) | <0.001 | 28,351 (89.6) | 13,132 (96.1) | <0.001 |
| ACEi/ARB/ARNI | 7,965 (75.8) | 2,179 (100.0) | <0.001 | 3,591 (74.7) | 2,642 (91.3) | <0.001 |
| MRA | 2,787 (26.5) | 1,219 (55.9) | <0.001 | 1,204 (25.1) | 1,362 (47.1) | <0.001 |

Values are n (%) unless otherwise indicated. Percentages are among patients with available data. P values are from χ² tests for categorical variables and t tests for continuous variables.
ACEi: angiotensin-converting enzyme inhibitor; ARB: angiotensin receptor blocker; ARNI: angiotensin receptor-neprilysin inhibitor; BMI: body mass index; CABG: coronary artery bypass grafting; COPD: chronic obstructive pulmonary disease; eGFR: estimated glomerular filtration rate; HF: heart failure; MRA: mineralocorticoid receptor antagonist; PCI: percutaneous coronary intervention; SD: standard deviation; SGLT2i: sodium-glucose cotransporter-2 inhibitor; TIA: transient ischemic attack.

### Provider Characteristics and treatment patterns

Of 7,946 providers in the ARNI analysis, 51.5% were female and 74.5% were physicians (SGLT2i analysis, 5,996 providers: 50.5% and 75.9%). By specialty, 69.7% of ARNI providers were primary care, 19.1% cardiology, 6.7% nephrology, and 4.4% endocrinology. Provider characteristics are summarized in **Supplemental Table S1**. There were no differences in medication use rate over time by provider type (Physician vs. APP) or sex, but there were differences by specialty (**Figure 1**). For ARNI, patients seen by cardiology had the highest use. For SGLT2i, patients seen by cardiology or endocrinology had the highest use. Patients seen by nephrologists had the lowest rates of use of both agents. Use increased over time for all provider specialties.

**Figure 1:**
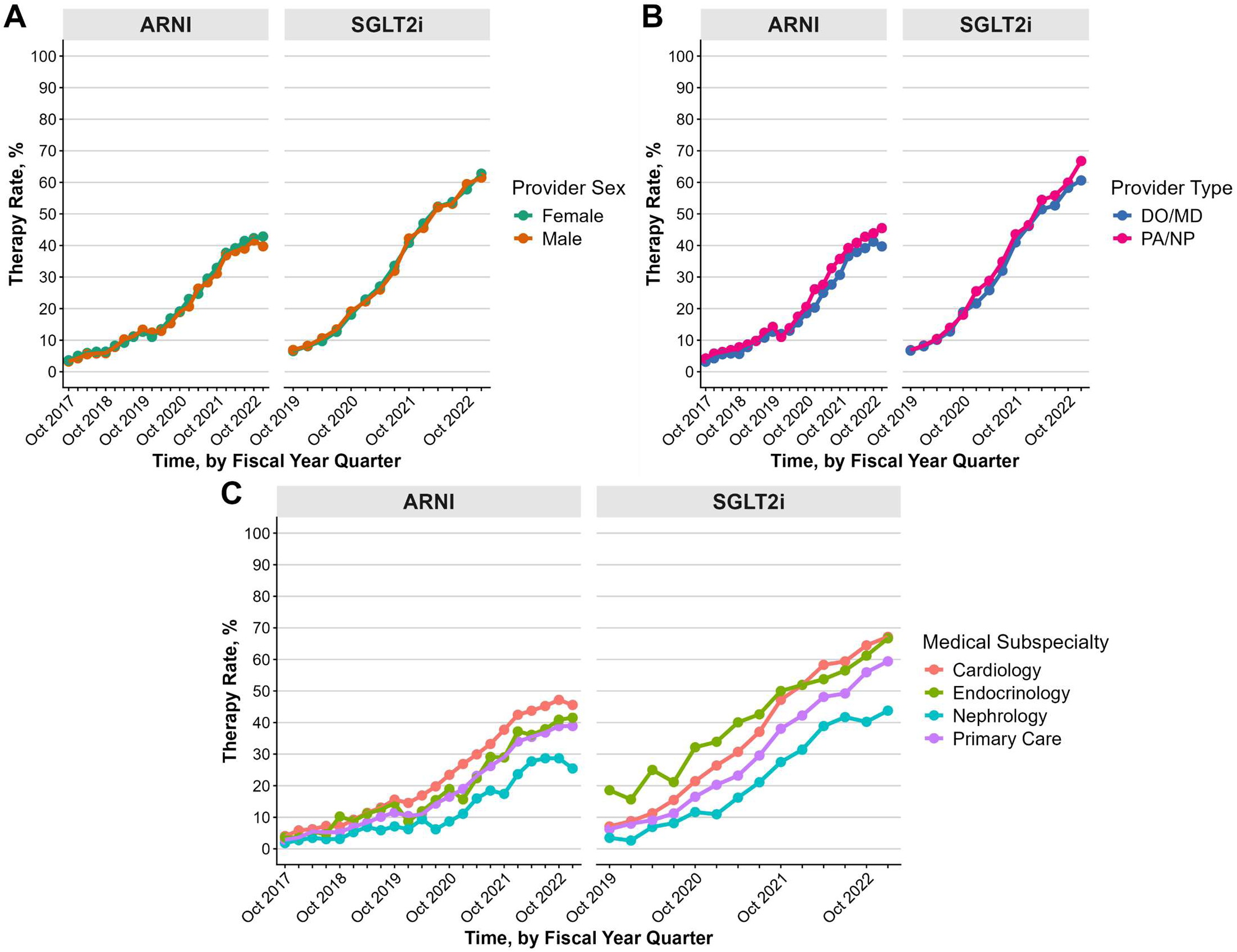
Medication utilization over time, by provider characteristics. Trends in mean medication utilization are shown over time by provider characteristics: physician, advance practice provider, or pharmacist provider type (Panel A), male or female sex (Panel B), and medical specialty (Panel C). Utilization increased for all categories throughout the study period. There were no substantial differences in trends over time by provider type or provider sex, but trends differed by specialty. HF: Heart Failure; ARNI: Angiotensin receptor-neprilysin inhibitor; SGLT2i: Sodium-glucose Cotransporter-2 Inhibitors

### Within-specialty variation in use

Within each clinical specialty, variability in the provider-level use of ARNI and SGLT2i was substantial (**Figure 2**). The distribution of ARNI use across providers was right-skewed: most providers had use below 50%, with a notable number in every specialty at near-zero use. For SGLT2i, the distribution was also right-skewed, with the largest number of providers at near-zero use in every specialty. For cardiology and endocrinology providers, there were also a greater number of providers with near-100% use.

**Figure 2:**
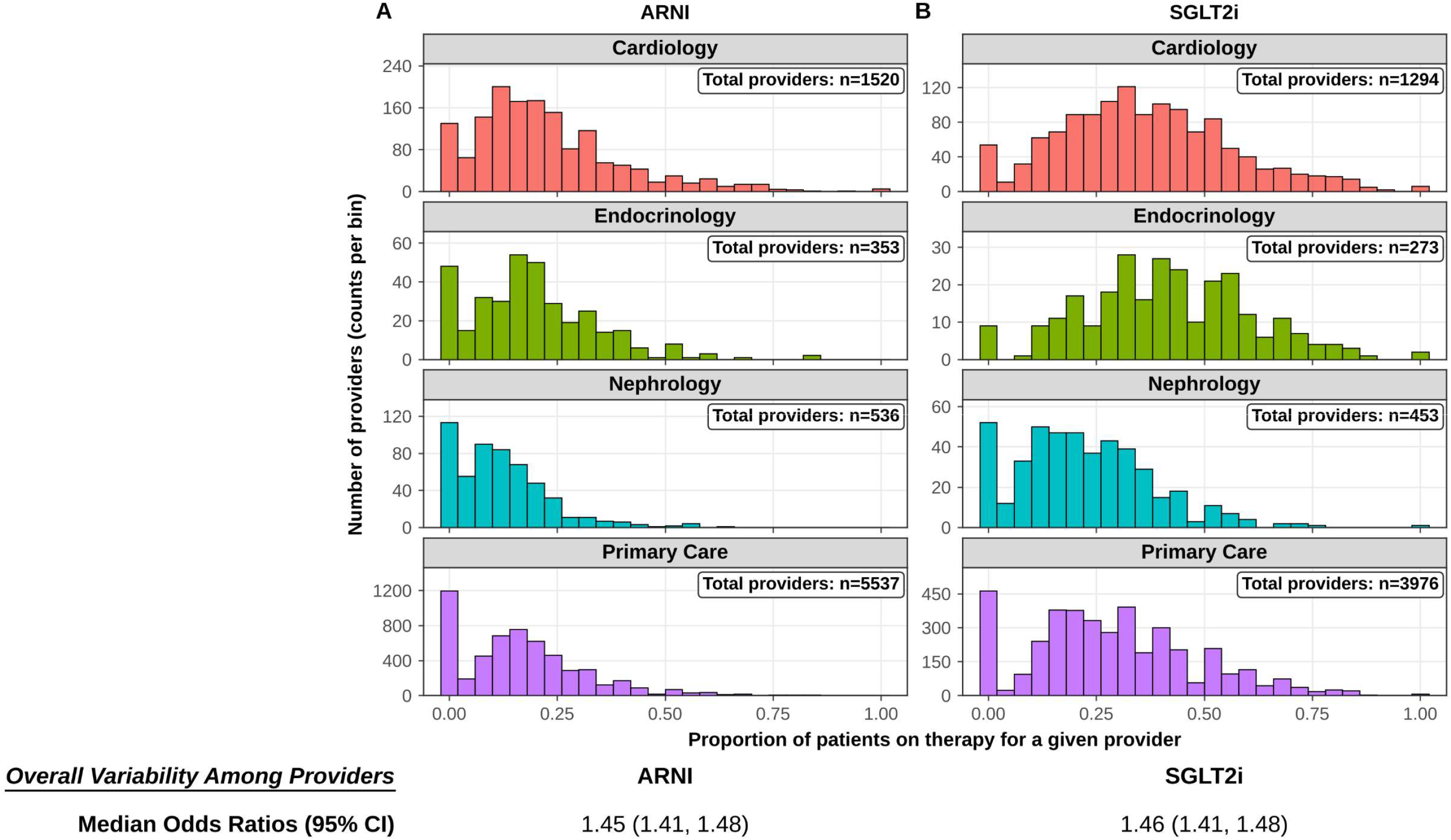
Within-specialty variability. Histograms show substantial variability in provider-level utilization of novel medications within each specialty. Overall median odds ratios (mORs) and breakdown of provider prescribing by specialty both show substantial variability in overall prescribing. Median odds ratios are adjusted for fiscal year quarter, patient age, gender, race, hypertension, renal function, type 2 diabetes, prior myocardial infarction, Veteran priority status, and VA site-level effects. HF: Heart Failure; ARNI: Angiotensin receptor-neprilysin inhibitor; SGLT2i: Sodium-glucose Cotransporter-2 Inhibitors

When examining the full cohort of providers, the median odds ratio (mOR) adjusted for patient characteristics was 1.74 (95% CI 1.65–1.99) for ARNI use and 1.82 (95% CI 1.69–2.11) for SGLT2i use.

When adding adjustment for site-level clustering, the mORs were 1.45 (95% CI 1.41-1.48) and 1.46 (95% CI 1.41-1.48) for ARNI and SGLT2i, respectively. In addition, the correlation between use of ARNI and use of SGLT2i at the provider level was modest (Pearson Correlation Coefficient 0.59), indicating providers with high use of one of the novel medications did not consistently have high use of the other.

Rate ratios varied widely within every specialty (**Table 2**). Across all providers, ARNI RRs were 0.46 at the 25th percentile, 1.62 at the 75th, and 2.92 at the 95th. For SGLT2i RRs were 0.58, 1.50, and 2.33, respectively. Across specialties, 95th percentile RRs ranged from 2.52 to 3.20 for ARNI and 1.82 to 2.50 for SGLT2i. RRs for the 25th percentile ranged from 0.37 to 0.56 for ARNI and 0.57 to 0.71 for SGLT2i. The 1st percentile use rate was 0% in every specialty for both drugs, and nephrology and primary care providers had rates of zero through at least the 10th percentile. The 99th percentile medication use rate in cardiology was 72.6% for ARNI and 85.8% for SGLT2i.

**Table 2:** Provider-level treatment rate ratios (RR) and prevalent use (% use) by percentile, unadjusted. Each cell shows the RR, defined as the provider’s treatment rate at that percentile divided by the median provider’s rate in the same column, followed by the provider’s rate (% use) in parentheses.

| **ARNI** | **All providers** | **Cardiology** | **Endocrinology** | **Primary care** | **Nephrology** |
|---|---|---|---|---|---|
| *Providers, N* | *N = 7,946* | *N = 1,520* | *N = 353* | *N = 5,537* | *N = 536* |
| 5th percentile | 0.00 (0.0%) | 0.00 (0.0%) | 0.00 (0.0%) | 0.00 (0.0%) | 0.00 (0.0%) |
| 25th percentile | 0.46 (7.1%) | 0.56 (10.8%) | 0.52 (9.1%) | 0.39 (5.9%) | 0.37 (3.8%) |
| **50th percentile (median; reference)** | **1.00 (15.4%)** | **1.00 (19.4%)** | **1.00 (17.6%)** | **1.00 (15.0%)** | **1.00 (10.3%)** |
| 75th percentile | 1.62 (25.0%) | 1.59 (30.8%) | 1.55 (27.3%) | 1.59 (23.8%) | 1.66 (17.1%) |
| 95th percentile | 2.92 (44.9%) | 2.99 (58.0%) | 2.52 (44.4%) | 2.86 (42.9%) | 3.20 (33.0%) |
| **SGLT2i** | **All providers** | **Cardiology** | **Endocrinology** | **Primary care** | **Nephrology** |
| *Providers, N* | *N = 5,996* | *N = 1,294* | *N = 273* | *N = 3,976* | *N = 453* |
| 5th percentile | 0.00 (0.0%) | 0.17 (6.0%) | 0.31 (12.5%) | 0.00 (0.0%) | 0.00 (0.0%) |
| 25th percentile | 0.58 (16.7%) | 0.64 (22.3%) | 0.71 (28.6%) | 0.61 (16.7%) | 0.57 (11.5%) |
| **50th percentile (median; reference)** | **1.00 (28.6%)** | **1.00 (35.0%)** | **1.00 (40.0%)** | **1.00 (27.3%)** | **1.00 (20.0%)** |
| 75th percentile | 1.50 (42.9%) | 1.39 (48.5%) | 1.38 (55.0%) | 1.47 (40.0%) | 1.56 (31.2%) |
| 95th percentile | 2.33 (66.7%) | 2.10 (73.3%) | 1.82 (72.9%) | 2.29 (62.5%) | 2.50 (50.0%) |

RR, rate ratio. % use = proportion of a provider’s qualifying encounters with the medication present. The median (50th percentile) provider has an RR of 1.00; an RR above 1 means a higher rate than the median provider in the same column, and 0.00 means a rate of zero. N is lower for SGLT2i because the analysis is restricted to encounters after the October 2019 DAPA-HF publication.
ARNI: angiotensin receptor-neprilysin inhibitor; SGLT2i: sodium-glucose cotransporter-2 inhibitor.

### Specialty and medication use

After adjustment for patient characteristics, copayment status, and site-level clustering, ARNI and SGLT2i varied significantly by provider specialty.(**Figure 3**). Patients who saw a cardiology provider (compared with those who did not see a cardiology provider) had the highest odds of both ARNI (OR 2.30 [95% CI 2.18-2.44]) and SGLT2i use (OR 1.99 [95% CI 1.88-2.11]). Patients who saw nephrology providers were less likely to be treated with ARNI (OR 0.67 [95% CI 0.63-0.72]) or SGLT2i (OR 0.64 [95% CI 0.59-0.69]). Visits with endocrinology and primary care were associated with increased SGLT2i use (OR 1.5 [95% CI 1.37-1.65]) and OR 1.12 [95% CI 1.05-1.19], respectively). In contrast, visits with endocrinology and primary care were not associated with increased ARNI use (OR 1.05 [95% CI 0.96- 1.14] and OR 1.05 [95% CI 1.00-1.11], respectively).

**Figure 3:**
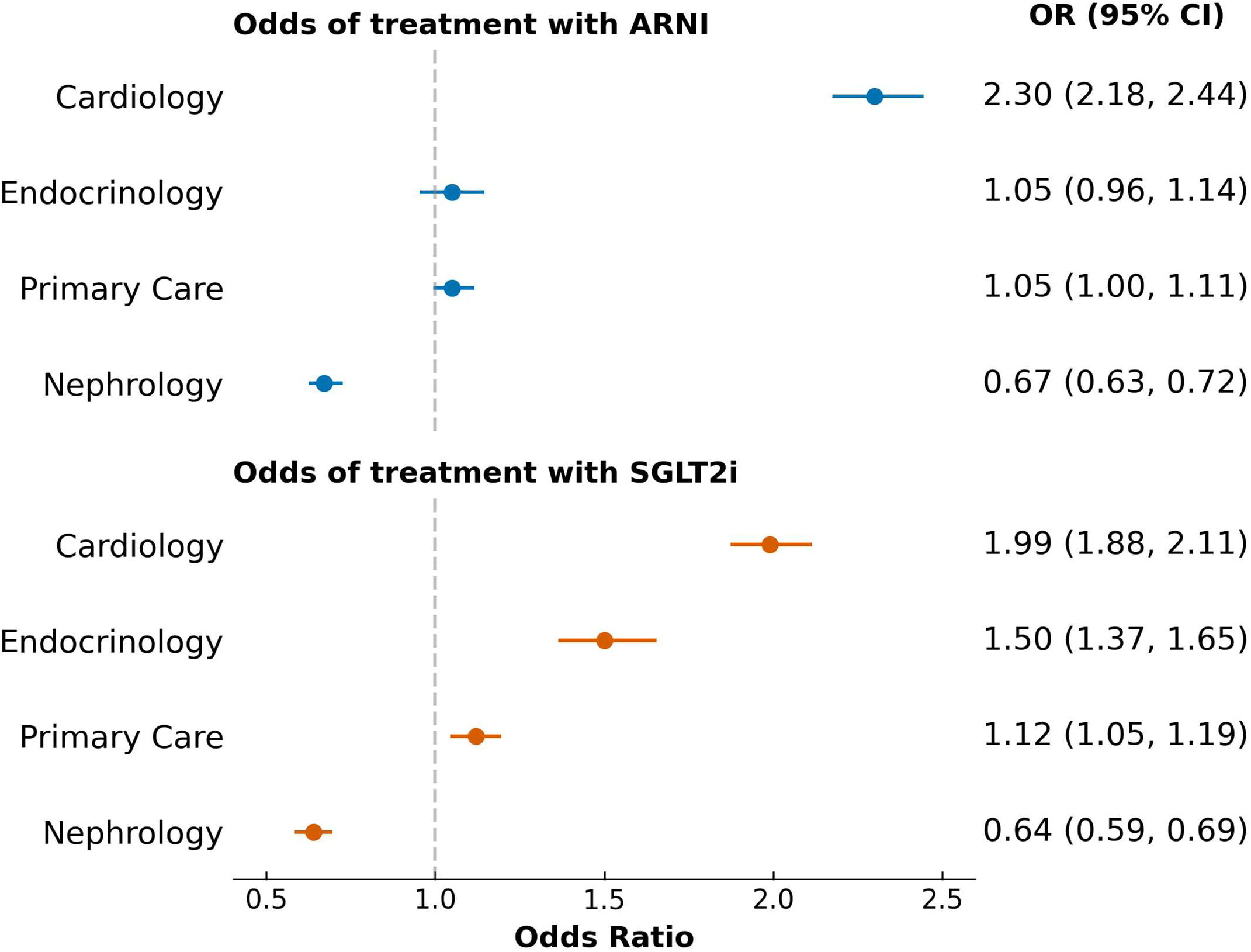
Forest Plot of Odds of receiving ARNI or SGLT2i treatment by specialty. Adjusted odds ratios (ORs) for receiving respective medication class treatment for having seen vs having not seen the indicated specialty. ORs are adjusted for: time of prescription (by fiscal year quarter) as well as patient age, sex, race, hypertension, renal function, history of prior myocardial infarction, type 2 diabetes, and Veteran priority status. HF: Heart Failure; ARNI: Angiotensin receptor-neprilysin inhibitor; SGLT2i: Sodium-glucose Cotransporter-2 Inhibitors

The association of cardiology and endocrinology visits with SGLT2i use changed over time, with the increased use associated with endocrinology visits waning in later years while the association of cardiology visits with use increased from 2019 to 2021 (**Supplemental Table S2**).

### Pharmacist visits

Over half of patients had at least one pharmacy visit within 6 months of hospitalization (55.0% and 55.1% of the ARNI and SGLT2i cohorts). When examined as a separate exposure, a pharmacist visit was independently associated with increased use of ARNI (OR 1.14 [95% CI 1.06-1.23]) and SGLT2i (OR 1.15 [95% CI 1.06-1.24]) after adjustment for patient characteristics and site-level clustering. Interaction testing did not show significant effect modification by specialty. However, the interaction term for cardiology and pharmacist visits demonstrated positive associations that neared statistical significance for ARNI (OR 1.08 [95% CI 0.97-1.19, p=0.164]) and SGLT2i (OR 1.11 [95% CI 1.00-1.23, p=0.052]) (Supplemental Table S4).

### Adoption after guideline incorporation

**Figure 4** shows post-hospitalization encounters by specialty and treatment status. In the full cohort, cardiology had the largest number of encounters (n = 95,489; 47.0% of total), followed by primary care (n = 86,728, 42.7%), nephrology (n = 14,777, 7.3%), and endocrinology (n = 6,093, 3.0%). Visits with patients who were not on therapy were each a potential opportunity to prescribe the indicated medication. Visits by cardiology providers had the highest proportion of new ARNI starts (11.8%; n=11,224, the greatest number) and the lowest proportion untreated at 6 months (78.7%). For SGLT2i, visits by endocrinology providers had the lowest proportion untreated at 6 months (58.1%), followed by cardiology (63.2%), and cardiology had the greatest number of new starts (n=11,027).

**Figure 4:**
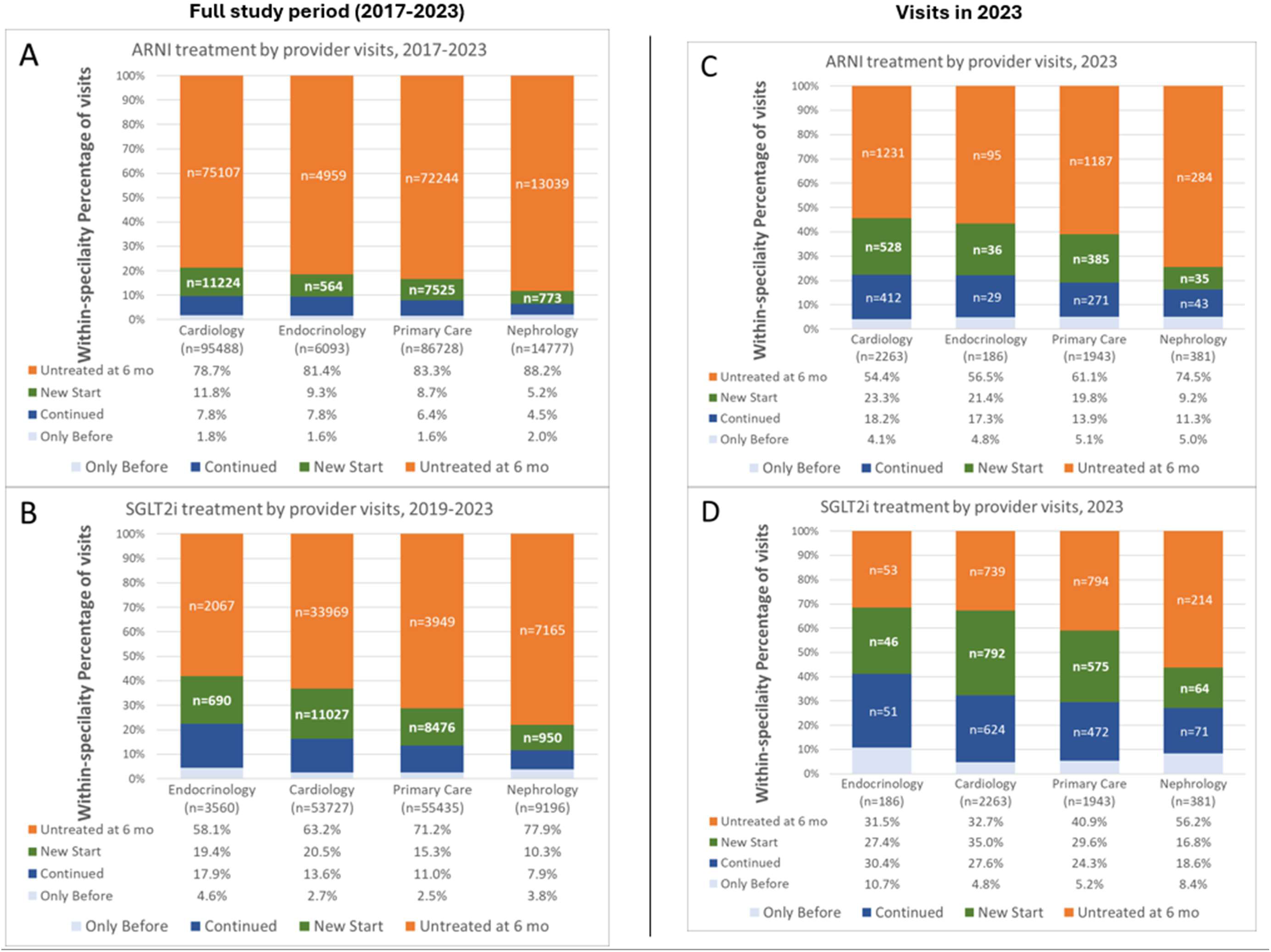
Novel medication treatment status by patient-provider visits, categorized by specialty. The proportion of provider visits seeing a patient who had medication treatment in 4 categories with respect to discharge: 1) only before; 2) continued; 3) new start; or 4) no treatment within 6 months of discharge. Panels separately show provider visits by specialty for ARNI and SGLT2i for the entire study period (**Panels A and B**, respectively) as well limited to the visits in 2023 (**Panels C and D**, respectively) HF: Heart Failure; ARNI: Angiotensin receptor-neprilysin inhibitor; SGLT2i: Sodium-glucose Cotransporter-2 Inhibitors

Because prevalent use credits providers for therapy begun by others, we also examined new starts, defined as no fill in the 6 months before admission and a fill at discharge or within 6 months afterward (**Table S3**). Across the study period, the new-start proportion of visits was 9.9% for ARNI and 17.3% for SGLT2i (after DAPA-HF). New starts rose from 2.6% in FY18 to 20.7% in FY23 for ARNI and from 5.0% in FY20 to 29.5% in FY23 for SGLT2i. Cardiology contributed the largest number of new starts for both ARNI (n=11,224) and SGLT2i (n=11,027).

In the final quarter of the study (January-March 2023), 58.4% of visits after HF hospitalization were with patients not treated with ARNI and 40.2% with patients not treated with SGLT2i, each a potential opportunity to prescribe the indicated therapy. Patients with a cardiology or endocrinology visit were least likely to remain untreated with ARNI or SGLT2i compared with other specialties. New starts of both medications were most common among patients with cardiology visits (**Figure 4**).

## DISCUSSION

In this retrospective cohort of Veterans hospitalized for HFrEF, which included over 7,900 providers and over 210,000 encounters, provider-level variation in ARNI and SGLT2i use was large within every specialty and independent of site effects. Use was also higher among patients seen by cardiology, endocrinology, and pharmacy providers within 6 months of discharge. Although use of both classes increased markedly over time, in early 2023 58.4% of visits after HF hospitalization were with patients not treated with ARNI and 40.2% with patients not treated with SGLT2i. These findings support an emphasis on specialty care, particularly cardiology, with adjunctive pharmacist follow-up after HF admission, and indicate that individual practice patterns within every specialty are an important potential target for implementation of GDMT.

Rapid initiation and up-titration of quadruple therapy is safe and effective, and site-level variation in ARNI and SGLT2i use has been described.^7,26^ STRONG-HF achieved high rates of GDMT through frequent post-discharge visits with interdisciplinary care teams,^19^ but the optimal composition of those teams is unknown, including whether follow-up with a cardiologist is required. Our study examines a period of rapidly increasing use of these medications after guideline incorporation in a national health system with comparatively low financial barriers to medication use.

### Provider-level variation

To our knowledge, this is the first characterization of provider-level variability in ARNI and SGLT2i use after HF hospitalization. Provider-level median odds ratios (mORs) were similar to prior reports of site-level variation,^25–27^ suggesting that which provider a patient sees may be as important as which site in the odds of treatment with ARNI and SGLT2i. The mOR, adjusted for site-level clustering, indicated a 45-46% difference in the odds of an ARNI or SGLT2i prescription from two randomly chosen providers. Provider-level variation therefore contributes in addition to site-level variation.

The empirical rate ratios describe probability at different percentiles of provider-level medication use relative to the median provider. Median use was highest for ARNI in cardiology (19.4%) and for SGLT2i in endocrinology (40.0%), followed by cardiology (35.0%). Across all providers, the rate ratio was 0.46 at the 25th percentile and 1.62 at the 75th, and a provider at the 95th percentile prescribed ARNI at 2.9 times the median rate; SGLT2i showed a similar pattern and magnitude (0.58, 1.50, and 2.33). In cardiology, the 95th percentile ARNI rate was nearly three times the median (rate ratio 2.99) and the 75th percentile was 1.59 times it. Across specialties, 95th percentile rate ratios ranged from 1.8 to 3.2. These unadjusted rates do not account for case mix, but the pattern was consistent across all specialties.

The distributions of use likely reflect individual implementation patterns for novel therapies over time. Some providers were early adopters with near-100% use, others were late- or non-adopters, and most fell in between, with right-skewed distributions (most providers at low use). ARNI was recommended in guidelines starting in 2016^3^ following key trial publication in 2014,^1^ and became increasingly favored over angiotensin-converting enzyme inhibitors or angiotensin receptor blockers, a shift reflected in the 2022 guidelines.^32^ SGLT2i was still early in use for HFrEF during the study period, with key trial publication for heart failure without diabetes in 2019 and guideline incorporation in 2021- 2022.^32,33^

Use of the two medication classes was only moderately correlated, suggesting that interventions may need to address ARNI and SGLT2i individually. ARNI and SGLT2i use rose steadily and exceeded reported utilization in non-VA cohorts,^8,13,22,32,33^ and the relative ease of access and low copayments in the VA may promote more comprehensive use. Provider-level variation may be even higher outside VA, where national standardization is lower.

We speculate that plausible drivers of this variation may include therapeutic inertia, differing familiarity with contemporary heart failure evidence, clinic workflow and time constraints, access to multidisciplinary heart failure infrastructure, availability of clinical decision support, and pharmacist collaboration. Our data cannot distinguish among them, and identifying them is an important target for future research.

### Post-HF hospitalization care by specialty and provider type

We found that most outpatient care after HF hospitalization was delivered by primary care and cardiology providers, together comprising 90% of post-HF hospitalization encounters. Importantly, we did not see substantial differences in ARNI or SGLT2i use by provider type (physician vs. APP). In VA, APPs commonly provide primary and specialty care with a broad scope of independent practice, which includes outpatient cardiology care and heart failure clinics. Our data support this care model.

In addition, over half of patients (55%) received a pharmacy visit within 6 months of hospital discharge. Clinical pharmacy encounters have been associated with increased prescription of SGLT2i and GLP-1 RA.^34–36^ In our study, pharmacist visits were associated with approximately 15% greater odds of treatment with ARNI and SGLT2i, regardless of which other specialties treated the patient. These results support further investigation on the role of pharmacists as independent or adjunctive providers after HF discharge. Our findings may be confounded by referral bias, i.e., patients already prescribed these medications by other providers may be more likely to be referred for pharmacy follow-up. VA clinical pharmacist practitioners practice under facility-level scopes of practice that include prescriptive authority,^29^ whereas outside VA, pharmacist prescribing generally depends on state legislation and collaborative practice agreements that are often narrow in practice, so these associations may not be directly generalizable outside VA.

Our results support a continuing role for specialty care after HF hospitalization, which is previously described to improve outcomes.^37,38^ A cardiology visit within 6 months of discharge was strongly associated with both ARNI and SGLT2i use, and the association persisted over time as overall use rose. An endocrinology visit was strongly associated with SGLT2i use early in the cohort (2019, immediately after DAPA-HF, when the class was already established for diabetes) but less so in later years. Cardiology and endocrinology visits were more likely to result in new medication starts, although endocrinology providers delivered a much smaller share of care. Lower use among patients with nephrology visits is an expected finding, because providers are more hesitant to prescribe these agents at the levels of kidney disease seen in nephrology practice, where benefit is less well established or therapy is considered contraindicated.

### Limitations

Our findings should be interpreted in the context of several limitations. First, we did not characterize which providers initiated or refilled prescriptions. We used a broad definition of medication use in which providers received credit even for continuing a medication started by a colleague; this may overestimate the prescribing of some providers but reflects real-world multidisciplinary management and complements studies focused on prescribing behavior.^8,22,39,40^ Second, administrative data and billing codes may misclassify patients. We did not assess ejection fraction, so the cohort may include patients with recovered or mid-range ejection fraction, although identification of systolic heart failure by diagnosis codes has been validated^27^ and indications have expanded over time. Medication use was based on VA pharmacy fills, and we cannot assess shared decision making or medications recommended but not filled. Third, this observational study cannot exclude unmeasured confounding; patients more likely to receive ARNI and SGLT2i may also be more likely to receive specialty care and pharmacist referral. Finally, the empirical rates are unadjusted for case mix and include providers with as few as five eligible encounters, so the extremes partly reflect sampling variability; the adjusted mOR addresses case mix but cannot exclude residual confounding.

### CONCLUSIONS

Use of ARNI and SGLT2i for patients with HF has increased over time in the VA, but provider- level variability was substantial despite a universal formulary and low out-of-pocket cost. Within every specialty, providers ranged from near-zero to near-100% use, and providers at the 95th percentile prescribed at 1.8 to 3.2 times the rate of the median provider of the same specialty. Use differed by specialty but not by provider sex or training, and patients seen by cardiology and endocrinology providers were most likely to be treated.

## Nonstandard Abbreviations and Acronyms

APP: advance practice provider
ARNI: angiotensin receptor-neprilysin inhibitor
CDW: Corporate Data Warehouse
CI: confidence interval
CKD: chronic kidney disease
COMIRB: Colorado Multiple Institutional Review Board
eGFR: estimated glomerular filtration rate
FY: fiscal year
GDMT: guideline-directed medical therapy
GLP-1 RA: glucagon-like peptide-1 receptor agonist
HbA1c: glycated hemoglobin
HF: heart failure
HFrEF: heart failure with reduced ejection fraction
mOR: median odds ratio
MRA: mineralocorticoid receptor antagonist
OR: odds ratio
RR: rate ratio
SGLT2i: sodium-glucose cotransporter-2 inhibitor
VA: Veterans Affairs

## Data Availability

VA data limited to VA system; data may be available upon reasonable request

